# Rapid spread of the SARS-CoV-2 δ variant in French regions in June 2021

**DOI:** 10.1101/2021.06.16.21259052

**Authors:** Samuel Alizon, Stéphanie Haim-Boukobza, Vincent Foulongne, Laura Verdurme, Sabine Trombert-Paolantoni, Emmanuel Lecorche, Bénédicte Roquebert, Mircea T. Sofonea

## Abstract

Analysing 9,030 variant-specific tests performed on SARS-CoV-2 positive samples collected in France between 31 May and 21 June 2021 reveals a rapid growth of the δ variant in 3 French regions. The next weeks will prove decisive but the magnitude of the estimated transmission advantages could represent a major challenge for public health authorities.

---

The evolution of SARS-CoV-2 variants has caused major epidemics in the United Kingdom (UK) [1, 2], Brazil [3], or South Africa [4]. In May 2021, the δ variant, which was first detected in India, was associated with an epidemic rebound in the UK [5]. According to Public Health England [6], this variant has a transmission advantage of approximately 66% (95% confidence interval (CI): [28,113]%) over the α variant. It is also slightly more prone to immune evasion [7]. As shown in 2021 with the emergence of the α variant, there is a two-month shift between the French and British epidemics [8, 9]. Therefore, it is timely to investigate the potential early spread of the δ variant to devise appropriate public health responses.

We analysed 9,030 RT-PCR variant-specific screening tests (VirSNiP assay, TIB Molbiol, Berlin, Germany) performed on samples collected in France between 31 May and 21 June 2021 that were tested positive for SARS-CoV-2. The assay screens for the presence of the E484K, the E484Q, and the L425R mutations (Supplementary Methods). A 484K+/484Q-/452R-profile is consistent with the B.1.351, P.1, and B.1.525 Pango lineages (i.e. β, γ, and η variants), whereas a 484K-/484Q-/452R+ profile is consistent with the B.1.617.2 lineage (i.e. δ variant) [10]. These tests originated from different French regions, with a dominance of the Île-de-France (the Paris area) (4,483/9,030 tests, i.e. 50%, Figure S1), and were mostly from non-hospitalised individuals (8,404/9,030 tests, i.e. 93%) (Supplementary Table S1).

## Variant-screening test results

Using a multinomial regression model as in [11], we found that samples bearing the L452R mutation and neither the E484K nor the E484Q mutation, i.e. consistent with the δ variant, tended to increase in the Île-de-France, Hauts-de-France, Normandie, and Provence-Alpes-Côte d’Azur regions compared to our reference, i.e. samples without any of the three mutations and consistent with the α variant (Table 1). Samples collected outside hospital settings were less likely to originate from β/γ/η variants than from the α variant. Finally, in the Hauts-de-France and Île-de-France regions, we found a temporal increase of β/γ/η variants compared to the α variant. In most regions, we found a temporal increase of ‘other’ test results compared to the α variant. This trend is more difficult to interpret since these tests may correspond to any variant.

**Table 1:** Factors associated with the detection of potential SARS-CoV-2 variants, as assessed by relative risk ratios (RRR) using a multinomial log-linear model, France, 31 May-21 June 2021 (n=8,190). In the model, screening tests consistent with an α variant are the reference for the RRR. ‘date:region’ indicates a temporal trend, i.e. an interaction between the date and the sampling region. See [11] for details about the methods used.

|  | **$\beta/\gamma/\eta$-like** | **δ-like** | **other** |
| --- | --- | --- | --- |
| age | 1.07 [1.00,1.10] | NS [0.94,1.20] | NS [0.98,1.10] |
| location (non-hospital) | 0.62 [0.49,0.8] | NS [0.64,1.60] | NS [0.71,1.10] |
| date:Normandie | NS [0.97,1.40] | 1.72 [1.30,2.20] | 1.4 [1.20,1.60] |
| date:Hauts-de-France | 1.29 [1.00,1.60] | 2.0 [1.50,2.70] | 1.52 [1.30,1.80] |
| date:Ile-de-France | 1.19 [1.10,1.30] | 2.23 [1.90,2.60] | 1.50 [1.40,1.60] |
| date:PACA | NS [0.75,1.30] | 1.92 [1.40,2.70] | NS [0.92,1.30] |
| date:Centre-Val-de-Loire | NS [0.56,1.20] | NS [0.51,1.70] | 1.29 [1.00,1.70] |
| date:other_regions | NS [0.71,1.00] | NS [0.70,1.30] | 1.40 [1.20,1.60] |

## Transmission advantages

To further quantify the temporal trends, we performed a statistical analysis using a logistic growth model following previous studies [12, 1, 2, 8]and pairwise comparisons between the three main interpretations of the test results, i.e. consistent with infections by δ, β/γ/η, or α variants. Between May 31 and June 21, the δ variant was found to have a median transmission advantage over the α variant greater than 70% in Île-de-France, Normandie, and Hauts-de-France (Figure 1A). In these three regions, we also found a significant transmission advantage of the δ variant over the β/γ/η variants (Figure 1B). Finally, consistently with our previous results [11], we find that β/γ/η variants have a significant, but smaller, transmission advantage over the α variant in these regions (Figure 1C). Transmission advantages tend to decrease with time (Figure S3), which could be linked to a delay in collecting the cases or a change in selection pressures.

**Figure 1:**
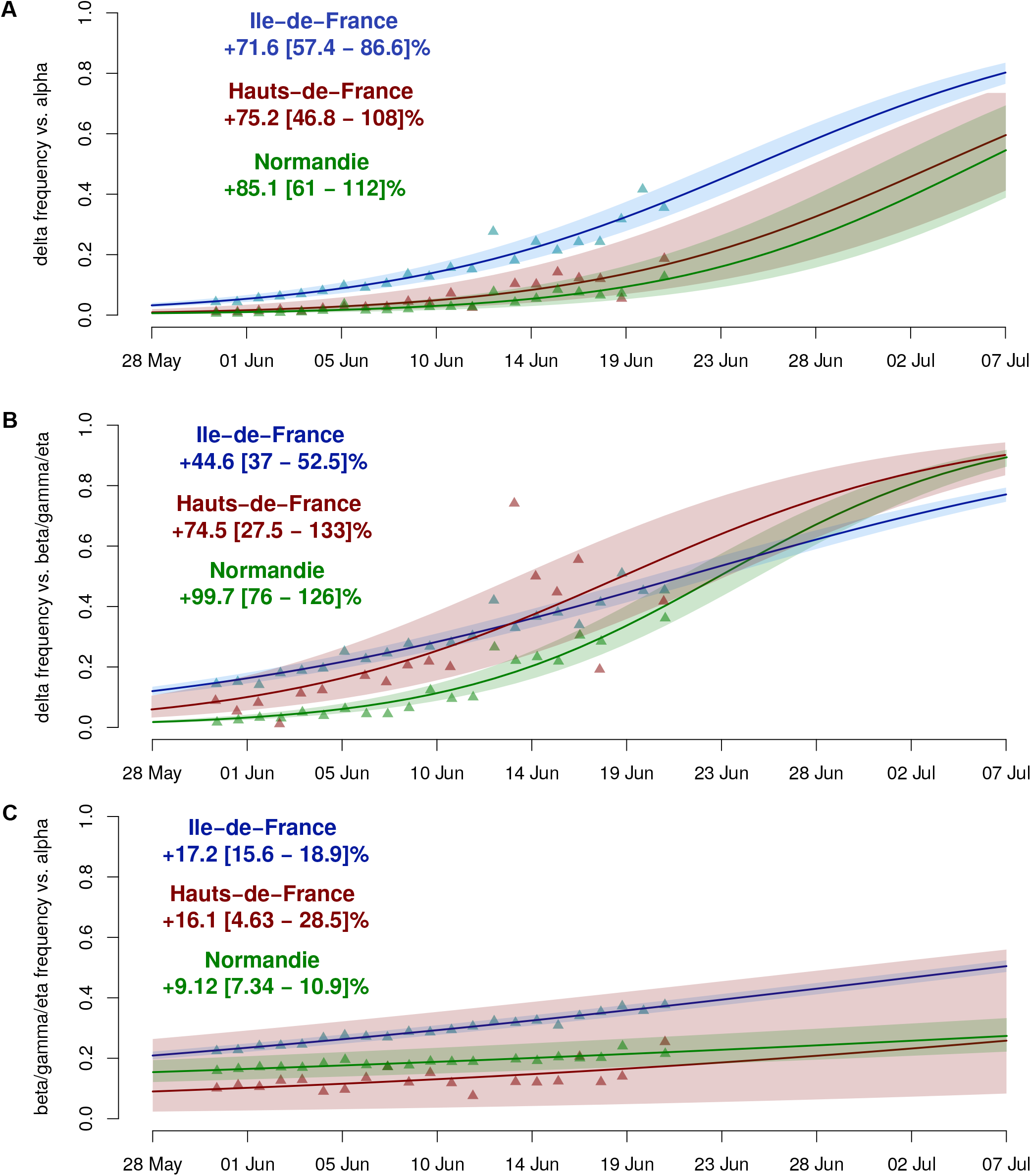
Estimated transmission advantage between variants in Ile-de-France, Hauts-de-France, and Normandie, 31 May-21 June 2021. A) δ variant versus the α variant, B) δ variant versus the β/γ/η variants, and C) the β/γ/η variants versus the α variant. The triangles indicate the generalised linear model (GLM)-fitted values and the line the output of the logistic growth model estimation. The shaded area represents the 95% CI. The estimated transmission advantages are shown in the panels.

## Doubling times

Using the incidence of δ variant consistent tests (Figure S4), we computed epidemic doubling times. When using a 14 days interval, we find low values such as 9.8 days (95CI: 6.3 – 22.6) in Île-de-France and 10.5 days (95CI: 6.9 – 21.9) in Hauts-de-France. However, in Île-de-France, this doubling time strongly increases with time and a 21 days interval yields a median doubling time of 26.9 days (Figure S5). This pattern could be explained by a delay in case reporting or, conversely, by an initial over-representation of superspreading events.

## Modelling epidemic scenarios

Using an epidemiological model of the French epidemic [13], we investigated the potential consequences of our results in the medium term (four months ahead). In Figure 2, we explored three scenarios (the A, B, and C stripes) that differ by arbitrary levels of vaccine rollout and back-to-school effect. In scenarios A and B, we assume no transmission increase (other than the transmission advantage of the δ variant) whereas in scenario C we impose a 10% *per capita* infectious contact rate increase at the start of the school year (September 1^st^) compared to June 2021. In scenarios A and C, the vaccine rollout is assumed to result in a 70% coverage of the whole population on September 1^st^, whereas in scenario B we consider a slower pace resulting in about 60% vaccine coverage by the end of the summer (see details in the Supplementary Methods). We find that the δ variant has the potential to initiate an epidemic rebound by the end of the summer that can be amplified by a slowdown in vaccine rollout and, even more, by an increase in infectious contact rate in September, which could lead to an Intensive Care Unit (ICU) overload (scenario C). Even in the absence of any additional mitigation, the residual COVID-19 hospital mortality for the second half of 2021 in scenarios A and B remains limited and corresponds to less than 10% of the current COVID-19 death toll in France. The trend of scenario C would however call for anticipation and control by the end of the summer.

**Figure 2:**
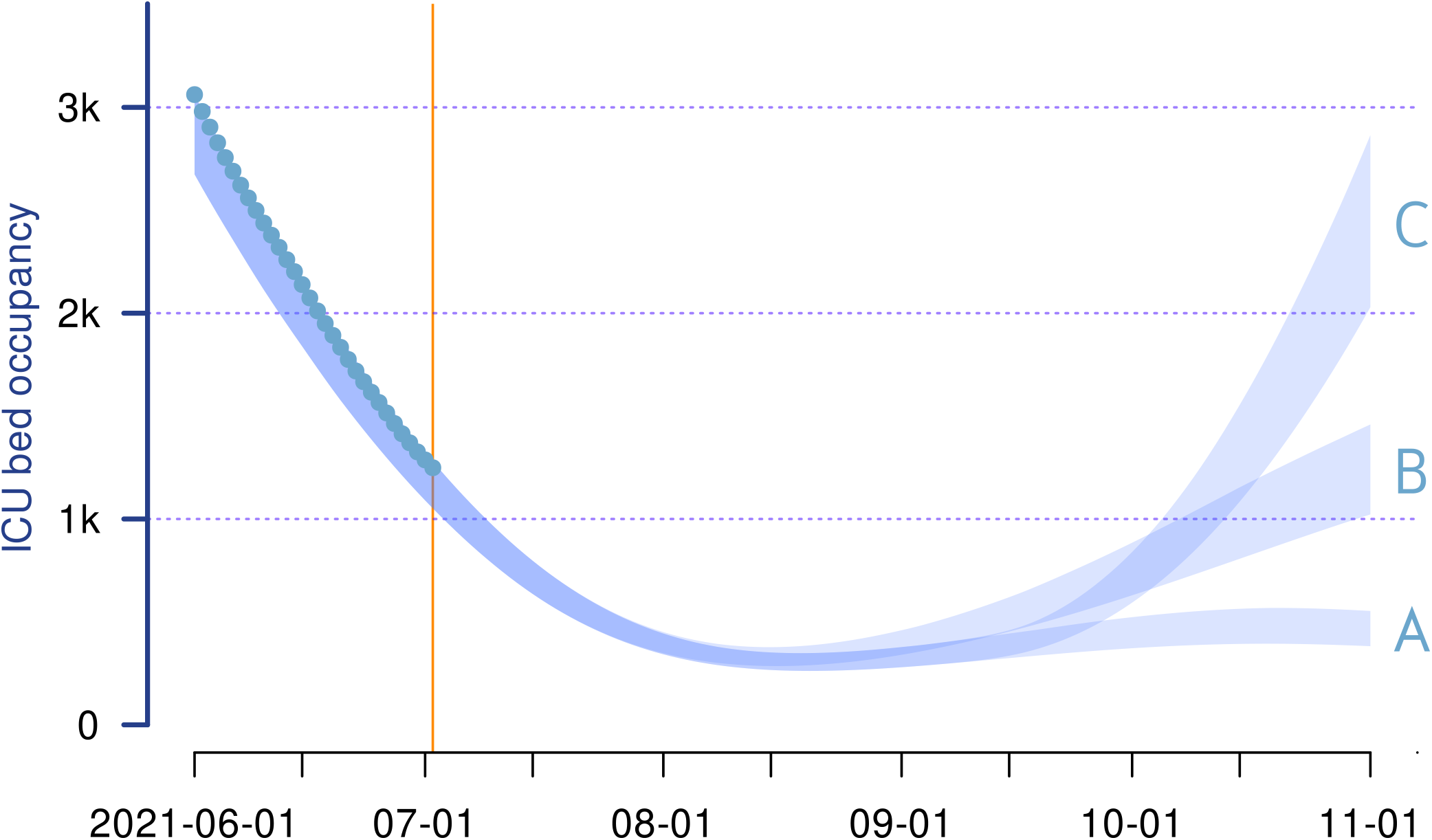
COVIDSIM projections of the French COVID-19 epidemic for different vaccine rollout and back-to-school effect scenarios. **A)** Fast vaccine rollout (i.e. 70% of the whole French population is assumed fully vaccinated on Sep 1^st^) and no back-to-school effect (i.e. the transmission increase is solely due to the spread of the δ variant, assumed to be 70% more transmissible than the α variant). **B)** Slower vaccine rollout (-10% coverage by the end of the summer compared to scenario A) and no back-to-school effect. **C)** Fast vaccine rollout and strong back-to-school effect (+10% increase in *per capita* infectious contact rate compared to June 2021). The blue stripes represent the current nationwide COVID-19 ICU bed occupancy according to the three scenarios. The width of each stripe corresponds to the range spanned by 95% of the associated simulations. The turquoise dots are the rolling 7-day averaged national COVID-19 ICU bed occupancy. The orange vertical line indicates the day the simulation was performed.

## Discussion

Screening for SARS-CoV-2 mutations bore by the δ variant on samples collected between 31 May and 21 June 2021 indicates that this variant might already be spreading rapidly in French regions. According to our estimates, the δ variant might already be more frequent than the α variant in Île-de-France by the end of June 2021.

There are several potential biases to this analysis, which we attempted to correct. First, since our dataset gathers results from local laboratories, there can be delays in data centralisation. To address this, we only analysed the data up to June 21 and ignored the data collected between June 22 and 28. Another potential issue has to do with the specificity of the test used. Our estimated frequency of δ variants is in the order of 4% of the positive tests on June 8. This is consistent with the outcome of national sequencing studies (0.2% on 11 May 2021 [14]) and doubling times comparable to that we estimated in the early stages of the δ variant epidemic in Île-de-France (Figure S5). Regarding the sampling scheme, the French authorities have announced strict monitoring of the spread of the δ variant, which could artificially enrich the data in δ variant positive tests. We do not know the context in which most tests were performed, except for that in a hospital setting, mainly for hospitalised patients. We expect hospital samples to be less impacted by a potential bias related to contact tracing because, aside from nosocomial infections, the contacts of a hospitalised person are unlikely to be hospitalised. Indeed, we find no association between the hospitalisation status and the δ variant in the multinomial model.

The estimated transmission advantages of the δ variant over the α variant are in line with that estimated in the UK [6] and from the contributions to the GISAID database [16]. Consistently with our earlier findings from April 2021 [11], we also find β/γ/η variants to have a transmission advantage over the α variant.

These results have major public health implications because they imply that variant δ could shift epidemic trends. In absence of specific interventions and based on the current vaccine rollout, our model tailored to the French epidemic suggests that this could cause another epidemic wave starting in August 2021. The magnitude and exact timing of this wave would depend on the exact transmission advantage of the variant, the increase in vaccination coverage and back-to-school effect as well as a potential loosening of social distancing and indoor mask-wearing. This picture could be further complicated by the simultaneous spread of variants β, γ, and δ.

## Data Availability

The data used for the analyses will be published along with the scripts upon acceptance.

## Acknowledgements

We thank Gilles Roullin, Eric Hedbaut and Virginie Dubois from CERBA for their technical assistance, and the ETE team from CNRS, IRD, and University of Montpellier for discussion.

## Ethical statement

This study has been approved by the Institutional Review Board of the CHU of Montpellier and is registered at ClinicalTrials.gov with identifier NCT04738331.

## Supplementary materials

### S1 Methods and data

We analyse the qualitative outcome of the SARS-CoV-2 VirSNiP E484Q/K and L452R assay from Tib Mol Biol (Berlin, Bermany) performed on samples that tested positive for SARS-CoV-2 in partner laboratories, mainly using partly the PerkinElmer SARS-CoV-2 Real-time RT-PCR Assay (Perkin Elmer).

The VirSNiP test targets three mutations specifically (E484K, E484Q, and L452R). Based on the genomics of the different lineages and their prevalence in France, the test results are interpreted as follows:

- E484K-/E484Q-/L452R-: potential infection by the *α* variant
- E484K-/E484Q-/L452R+: potential infection by the *δ* variant
- E484K+/E484Q-/L452R-: potential infection by the *β, γ*, or *η* variant

All the other combinations were rare and grouped in the “other” category (see below for the details).

The data was extracted on June 28, 2021 and the most recent test results were from June 26, 2021. However, to correct for potential delays in the recording of the results from local partners, we did not include any tests performed after June 21, 2021. Ve also only included from individuals from 5 to 80 years old, and included at most one sample per individual.

The statistical methods are described in details in 1J. The data and R script used will be provided upon publication.

### S2 Data characteristics

The characteristics of the data are shown in Table S1.

Among the other’ test results, the majority corresponded to uninterpretable test results (751/1394, i.e. 54%) and a large fraction to tests without the L452R mutant and an ambiguity for the E484 mutation (524/1394, i.e. 38%), which could be indicative of an infected by a *β, γ*, or *η* variant with a low virus load. Note that this assumption is conservative to avoid overestimating the spread of the *δ* variant. In fact, some of the tests in the other’ category (74/1394, i.e. 5%) find a clear signal for the L452R mutation with an ambiguity regarding the E484 mutation and could, therefore, be associated with this variant.

### S3 Epidemiological model

The epidemiological model used to simulate the scenarios shown in Figure 2 is based on the discrete-timeframework adjusted to the French epidemic the methodology of which is detailed in 2J. The original model was extended to take into account the vaccine rollout, the extrapolation of which was based the trend observed by the of June. Ve made the simplifying (and optimistic) assumption that the vaccine prevents 90% of critical infections and 80% of secondary infections directly from the first dose injection, while we assumed natural immunity to prevent 84% of reinfections 3J, regardless of the virus genotype.

The modelled spread of the *δ* variant was based on a initial 25%-frequency on Jun 29th and a 70% transmission advantage compared to the *α* variant, assumed itself to have a 40% transmission advantage with respect to the historical strain 1J.

**Table S1:** Characteristics of the samples analysed via the VirSNiP assay, France, 31 May-21 June 2021 (*n* = 9, 030).

| Characteristics |  | Value |
| --- | --- | --- |
| Age | *median [min,max]* | 34 [6,84] |
| Sample origin | general population | 8404 (93.1%) |
|  | hospital | 626 (6.9%) |
| Sampling date | *median [min,max]* | June 7 [May 31, June 21] |
| Region | Ile-de-France | 4483 (49.9%) |
|  | Normandie | 1534 (17.1%) |
|  | Hauts-de-France | 1040 (11.6%) |
|  | PACA | 518 (5.8%) |
|  | Centre-Val-de-Loire | 415 (4.6%) |
|  | other | 993 (11.0%) |
| Test result | α-like | 4478 (49.6%) |
| | $\beta/\gamma/\eta$-like | 1165 (12.9%) |
| | δ-like | 381 (4.2%) |
|  | other | 1235 (13.7%) |
|  | uninterpretable | 1771 (19.6%) |

## S4 Supplementary Results

**Figure S1:**
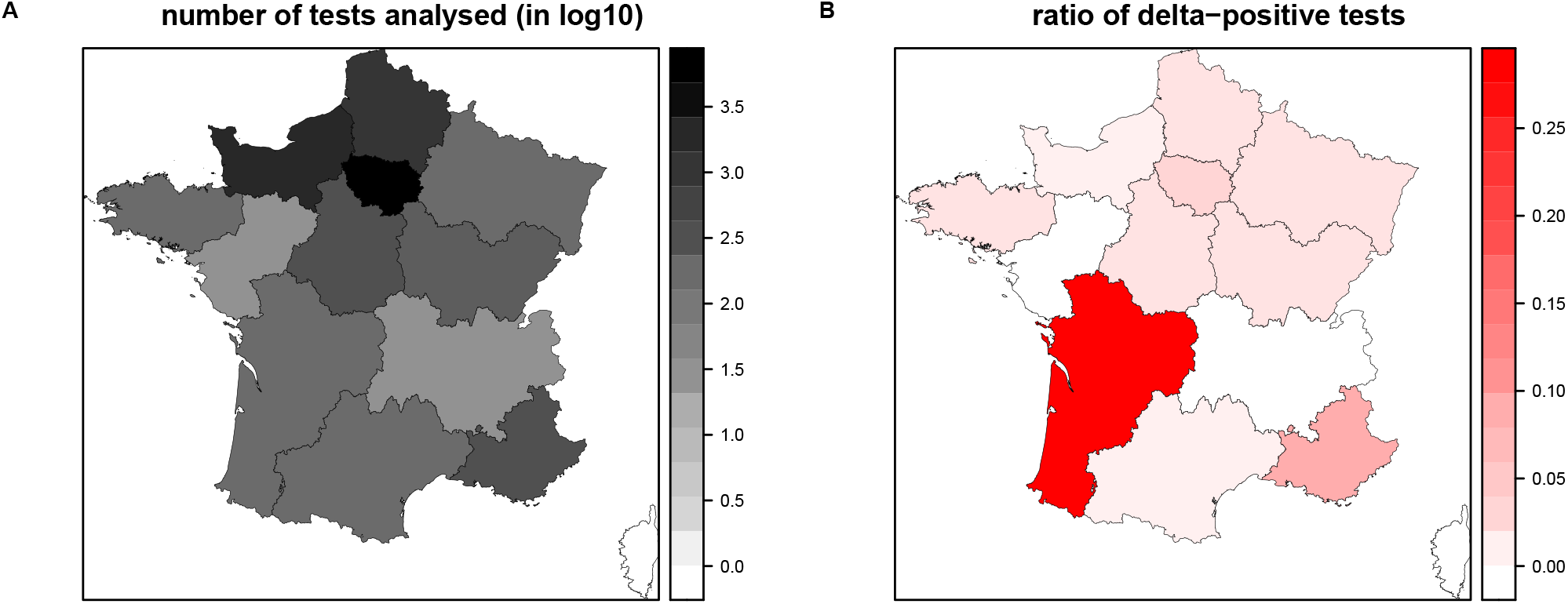
A) Number of tests analysed in French regions and B) Proportion of the tests consistent with an infection by the *δ* variant

**Figure S2:**
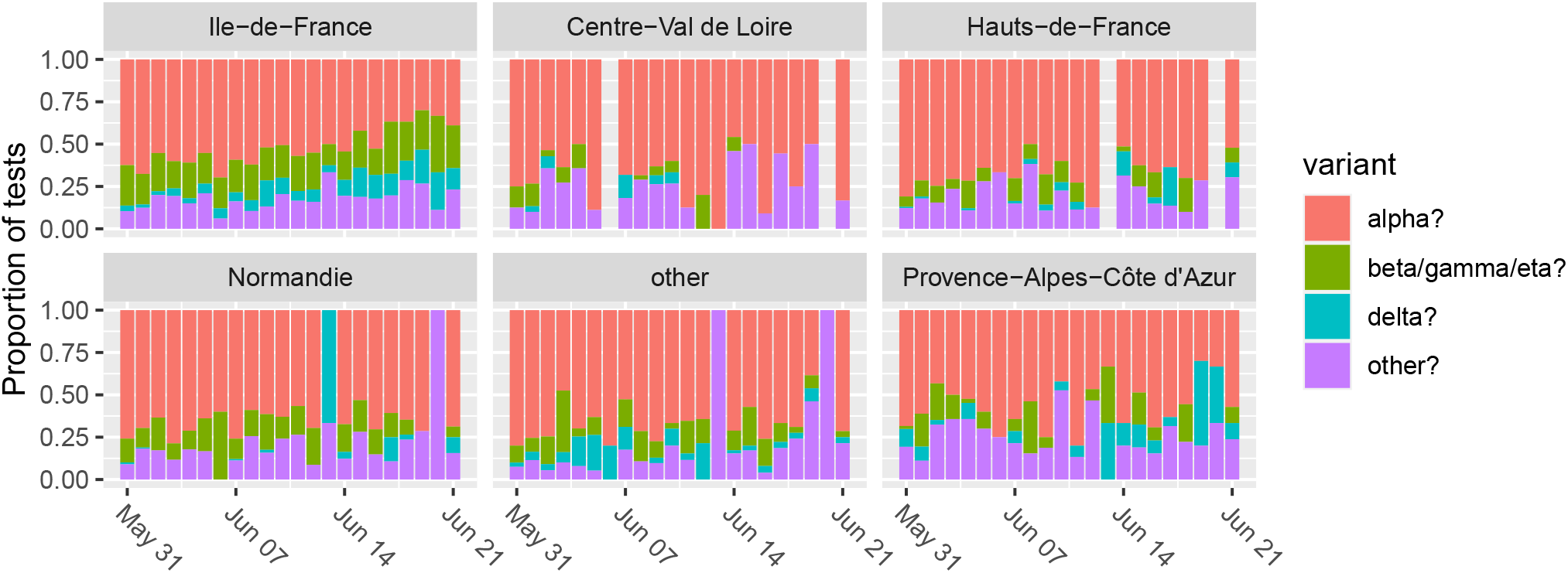
Proportion of tests associated with the main variants in the most densely sampled French regions.

**Figure S3:**
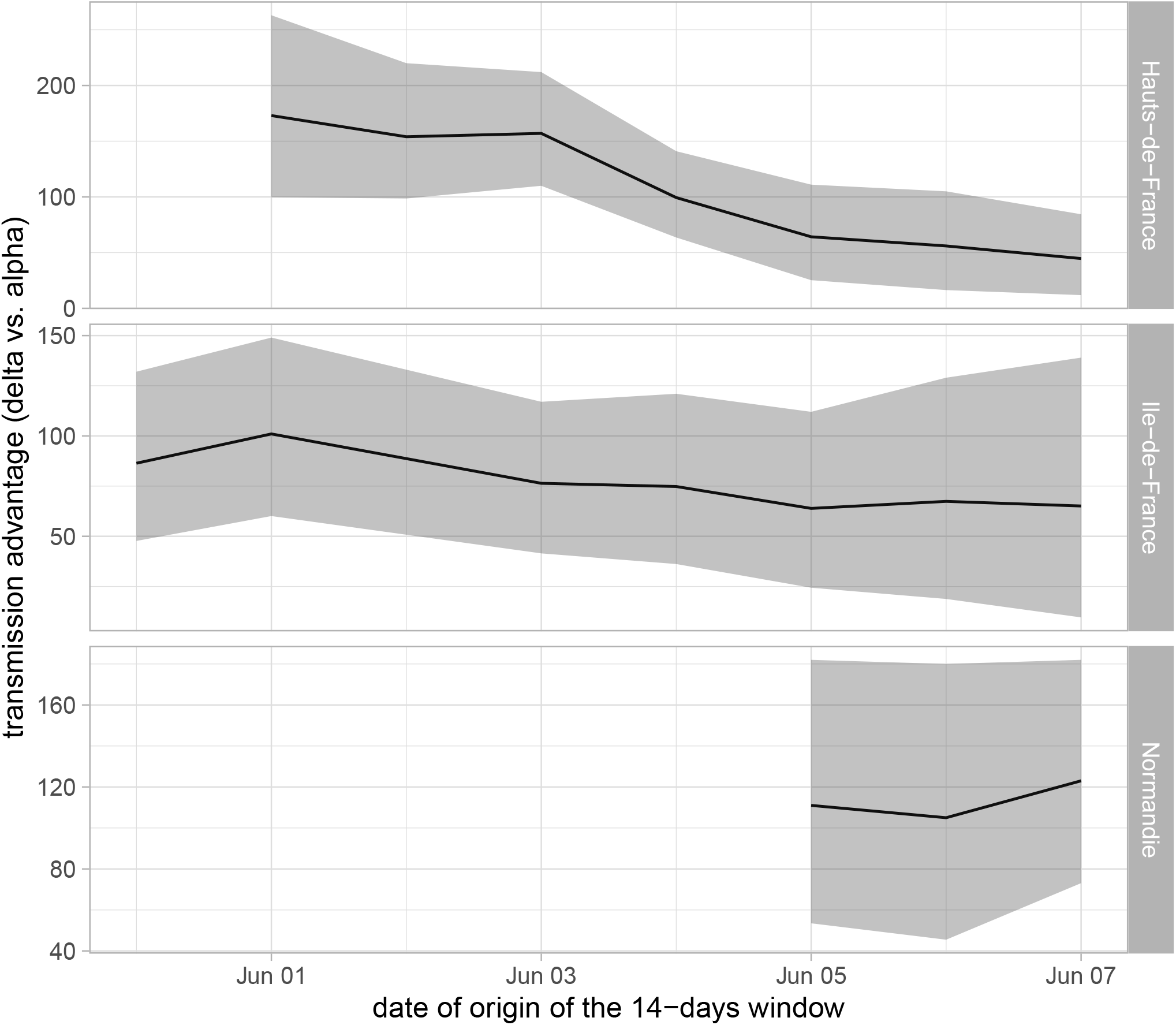
Sliding 14-days window estimating the transmission advantage of the *δ* variant of the *α* variant in 3 French regions.

**Figure S4:**
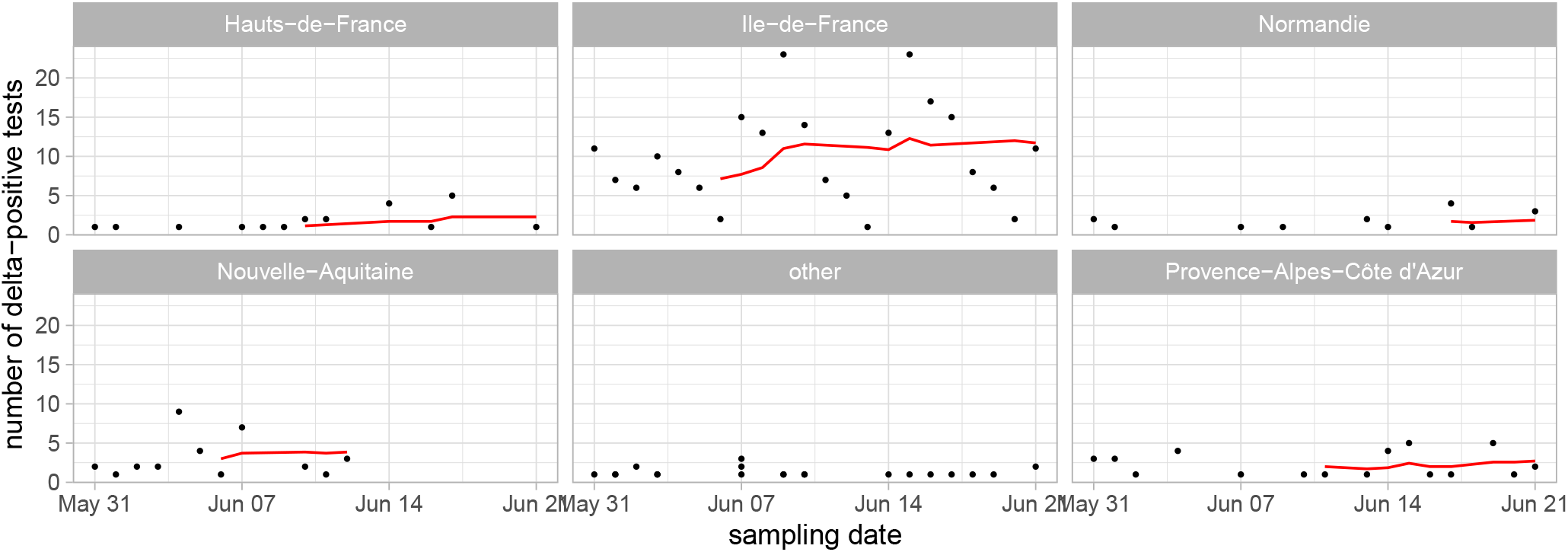
A) Number of tests consistent with an infection by the *δ* variant. The red line shows a right-aligned 7-day rolling mean.

**Figure S5:**
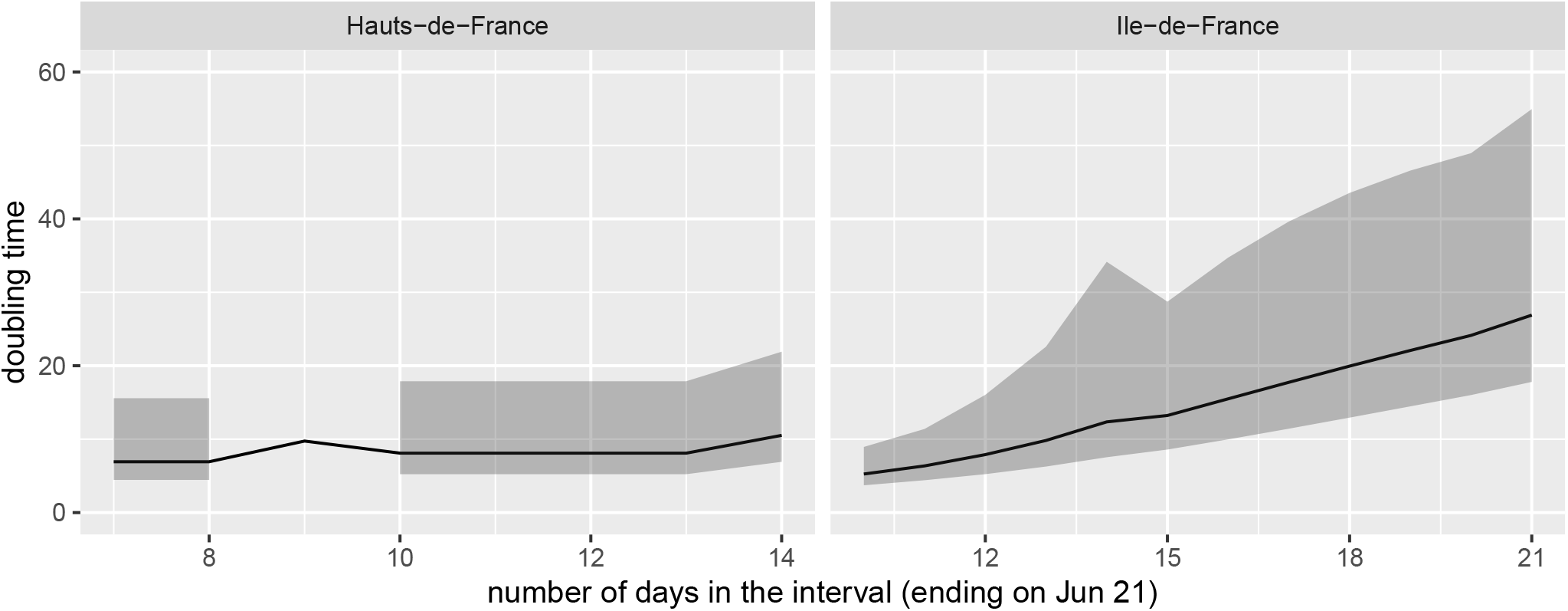
Epidemic doubling time as a function of the interval considered. The interval starts on May 31 in Ile-de-France and on June 7 in Hauts-de-France (due to a lack of data). The confidence interval are calculated using the regression model between the log of the number of cases and the time.

